# Deep brain stimulation-induced local evoked potentials outperform spectral features in spatial and clinical STN mapping

**DOI:** 10.1101/2025.06.14.25329308

**Authors:** Enrico Opri, Faical Isbaine, Seyyed Bahram Borgheai, Emily Bence, Roohollah Jafari Deligani, Jon T. Willie, Robert E. Gross, Nicholas Au Yong, Svjetlana Miocinovic

## Abstract

**Objective:** Deep brain stimulation (DBS) of the subthalamic nucleus (STN) is an established therapy for Parkinson’s Disease (PD). Yet, optimizing lead placement and stimulation programming remains challenging. Current techniques rely on imaging and intraoperative microelectrode recordings (MER), while programming rely on trial-and-error clinical testing, which can be time-consuming. DBS-induced local evoked potentials (DLEP), also known as evoked resonant neural activity (ERNA), have emerged as a potential alternative electrophysiological marker for mapping. However, direct comparisons with traditional spectral features, such as beta-band, high-frequency oscillations (HFOs), and aperiodic component have been lacking.

**Approach:** We evaluated DLEP across 39 STN DBS leads across 31 subjects with PD undergoing DBS surgery, using both a single-pulse and high-frequency burst stimulation paradigms. We developed a novel artifact-removal method to enable monopolar DLEP recovery, including estimating the DLEP amplitudes at stimulated contacts, further enhancing spatial sampling of DLEP. We evaluated spectral features and DLEP in respect to imaging-based and MER-based localization, and its predictive power for post-operative programming.

**Main Results:** DLEP showed great spatial consistency, maximizing within STN with 100% accuracy for single-pulse and 84.62% for burst stimulation, surpassing spectral measures including beta (89.74%) and HFO (82.05%). DLEP better correlated with clinical outcomes (single-pulses ρ=−0.33, high-frequency bursts ρ=−0.26), than spectral measures (beta ρ=−0.25, HFO ρ=0.05). Furthermore, single-pulses at low-frequencies are sufficient for DLEP-based mapping.

**Significance:** We show how DLEP provide higher STN-spatial specificity and correlation with postoperative programming compared to spectral features. To support clinical translation of DLEP, we developed two methods aimed to recover artifact-free DLEP and estimating DLEP amplitudes at stimulating contacts. DLEP appear distinct from beta and HFO activity, yet strongly tied to aperiodic spectral components, suggesting that DLEP amplitude reflects underlying STN excitability. This study highlights that DLEP are a robust and clinically valuable marker for DBS targeting and programming.

## 1. Introduction

Deep brain stimulation (DBS) of the subthalamic nucleus (STN) has become a standard therapy for patients with Parkinson’s disease (PD). This procedure includes surgical implantation of leads into the basal ganglia, enabling the delivery of electrical stimulation to modulate neural activity and alleviate symptoms. However, challenges persist in optimizing treatment outcomes [1, 2, 3, 4, 5], including complex surgical targeting and time-consuming postoperative selection of the most effective stimulation parameters. Typically, imaging-based surgical planning is refined intraoperatively through functional-mapping, which leverages micro-electrode recordings (MER) and intraoperative stimulation to achieve accurate placement of the DBS lead [6]. This is critical to maximize symptom relief while minimizing side effects induced from stimulation spread to unintended areas[7, 8]. However, this approach requires an expert neurophysiologist, is time-consuming, and can suffer from intra-rater subjectivity [6, 9, 10, 11].

The spatial distribution of beta-band power (13-35Hz)[4, 12] has been proposed as an alternative physiological marker for functional-mapping. However, it has notable limitations: large variability across patients in peak frequency/bandwidth[13], variability over time [14, 15, 16], peak absence in some patients [17], and requirement for patients to be awake, off dopaminergic medications, and remain at rest during recordings [16]. Postoperatively, during DBS programming, beta power has been proposed as a biomarker to assist in stimulation parameter selection, but similar limitations apply[18, 19]. This has led to a renewed interest in finding a marker that can reliably assist with physiological target mapping and programming.

DBS induced local evoked potentials (DLEP)[20, 21], also known as evoked resonant neural activity (ERNA) [22, 23, 24, 25, 26, 27, 28, 29], DBS evoked potential (DEP) [30], or generally evoked compound action potential (ECAP) [31], have emerged as a promising marker for intraoperative mapping. DLEP is elicited by stimulation in the target region and is typically recorded using inactive, adjacent DBS contacts (Fig. 1A). Prior studies suggest that the DLEP amplitude may be largest at the contacts within the sensorimotor STN and associated with the best therapeutic benefit [23, 25]. DLEP is also detectable within the globus pallidus internus (GPi), and it has been suggested that DLEP may result from activation of reciprocal connections between the external globus pallidus and STN [28, 31]. DLEP is attractive as they have 1-2 orders of magnitude larger amplitudes compared to STN/GPi local field potential activity[22], providing excellent signal-to-noise ratio (SNR) characteristics. However, in previous work DLEP typically required to be recorded with a bipolar montage, or biphasic symmetric stimulation to limit effects of stimulation artifacts [20, 22, 23, 26], limiting broader clinical applications.

**Figure 1.**
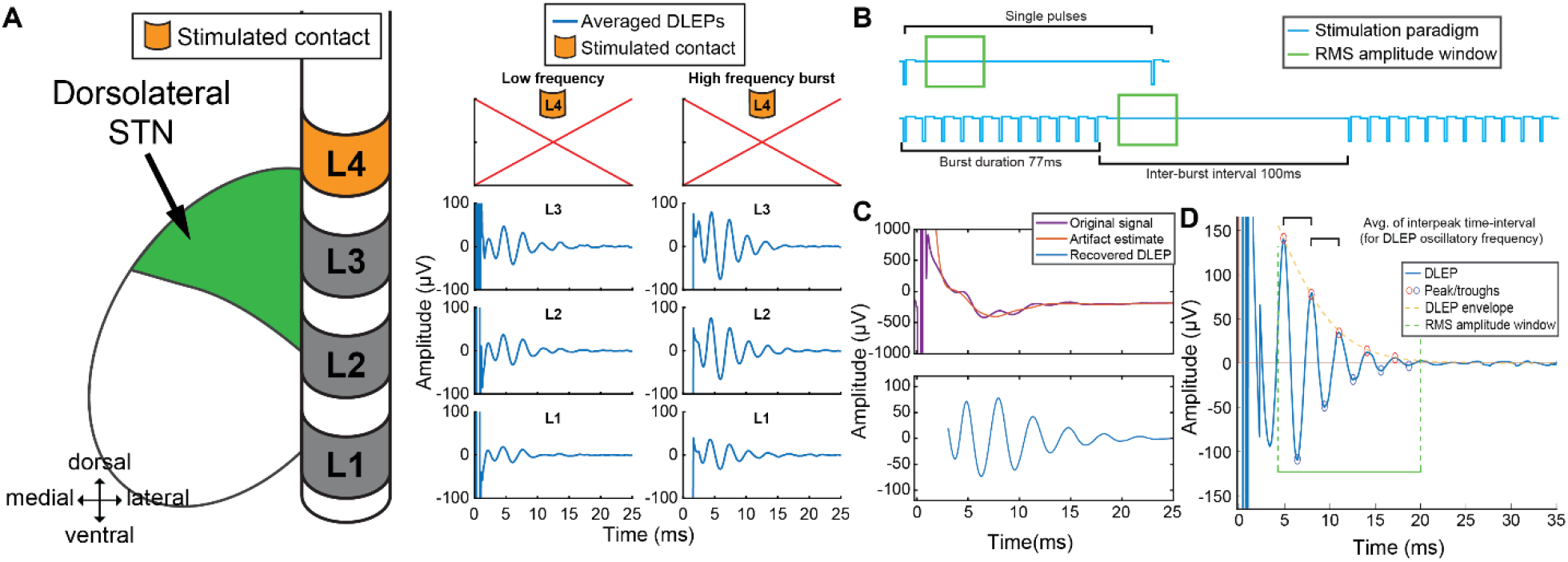
Stimulation protocol for recovery of DLEP and neural recruitment. (A) DLEP responses in single subject with PD stimulating contact on level 4 (L4) both at low frequency (LF-DLEP) and high frequency burst (HF-DLEP) stimulation; (B) Stimulation protocol included 1) low frequency stimulation at 10Hz (each pulse every 100ms) for 12 seconds with 3 seconds pause while cycling through each lead contact (in randomized order); 2) burst pattern of 11 stimulation pulses delivered at 130Hz with 100ms interval with no stimulation repeated for 30 times, with 3 seconds pause while cycling through each stimulated contact (randomized). Each stim In both cases stimulation was delivered with an asymmetric waveform, 60us pulse for first phase, and 480us for the second phase. (C) Artifact removal for DLEP recovery, from the STN of a PD patient. (D) Morphological descriptors of DLEP: RMS, oscillatory frequency (average interpeak time-interval), decay constant (estimated from exponential fit of the DLEP envelope). Features will be computed within the time-window specified in green (4-20ms).

The primary goal of this study was to determine whether DLEP can reliably localize sensorimotor STN and assess their potential as a marker for guiding DBS therapy compared with MER and beta power. We hypothesized that DLEP provide greater spatial specificity, greater consistency across individuals, and a stronger correlation with therapeutic outcomes than beta power. In order to answer these questions, we developed novel artifact-removal and signal processing methods to record DLEP with high spatial resolution. Furthermore, we investigated if there was information loss in spatial localization using different stimulation patterns, with burst of high-frequency (130Hz) stimulation or single pulses. This work seeks to understand the usefulness of neural-markers for optimizing DBS therapy in individuals affected by PD.

## 2. Method

### 2.1. Participants

Patients with idiopathic PD scheduled to undergo STN DBS surgery at Emory University were recruited for the study (Table 1). Written, informed consent was obtained before surgery under a protocol approved by the Institutional Review Board.

**Table 1.** Patient demographics and stimulation paradigm. Subject demographic, stimulation paradigm tested, and lead coordinates. Stimulation paradigm includes if the LF DLEP or HF DLEP protocol were run for that subject/hemisphere.

| PatID | Hemi | Age | Sex | Stimulation paradigm | DBS lead bottom contact MNI coordinates (mm) | DBS Lead Model |
| --- | --- | --- | --- | --- | --- | --- |
| S01 | R | 6X | M | LF | 12.80, -15.69, -7.63 | MDT 3389 |
| S02 | R | 5X | M | LF | 10.68, -12.52, -8.80 | MDT 3389 |
| S03 | L | 4X | M | LF | -13.77, -11.45, -8.40 | MDT 3389 |
| S04 | R | 5X | M | LF | 11.49, -13.93, -7.53 | MDT 3389 |
| S05 | L | 4X | M | LF | -11.15, -13.39, -11.31 | ABT 6172 |
| S06 | L | 4X | M | LF | -15.39, -13.67, -4.77 | ABT 6172 |
| S07 | R | 4X | M | LF | 11.47, -16.44, -9.57 | ABT 6172 |
| S08 | R | 4X | M | LF | 12.43, -14.01, -9.42 | ABT 6172 |
| S09 | R | 5X | M | LF | 11.19, -14.11, -10.08 | BSC 2202 |
| S10 | L | 6X | M | LF | -13.02, -14.96, -6.72 | ABT 6172 |
| S11 | L | 6X | M | LF and HF | -11.48, -13.90, -8.23 | ABT 6172 |
| S12 | R | 4X | M | LF | 10.97, -14.91, -9.33 | BSC 2202 |
| S13 | R | 6X | M | LF and HF | 11.71, -12.78, -9.95 | ABT 6172 |
| S14 | L | 6X | M | LF and HF | -11.21, -13.17, -8.16 | BSC 2202 |
| S15 | L | 6X | M | LF | -11.33, -13.57, -8.94 | BSC 2202 |
| S16 | L | 5X | F | LF | -10.21, -13.88, -8.84 | ABT 6172 |
|  | R |  |  | LF and HF | 11.15, -13.87, -8.93 | ABT 6172 |
| S17 | L | 5X | M | LF and HF | -14.50, -14.39, -7.88 | MDT B33005 |
|  | R |  |  | LF and HF | 11.55, -15.11, -8.98 | MDT B33005 |
| S18 | L | 4X | F | LF | -12.55, -13.41, -9.09 | BSC 2202 |
|  | R |  |  | LF | 10.83, -14.02, -9.91 | BSC 2202 |
| S19 | L | 6X | M | LF and HF | -10.46, -12.45, -8.13 | BSC 2202 |
| S20 | R | 6X | M | LF and HF | 10.81, -13.29, -9.05 | MDT B33005 |
| S21 | L | 8X | M | LF | -13.55, -16.55, -5.93 | BSC 2202 |
| S22 | R | 7X | M | LF and HF | 8.52, -12.93, -11.33 | BSC 2202 |
| S23 | L | 5X | M | LF and HF | -10.89, -14.14, -8.63 | BSC 2202 |
| S24 | R | 6X | M | LF and HF | 12.45, -13.64, -8.61 | BSC 2202 |
| S25 | R | 6X | M | LF and HF | 10.97, -14.15, -9.88 | BSC 2202 |
| S26 | L | 5X | F | LF | -12.57, -14.16, -8.70 | BSC 2202 |
|  | R |  |  | LF and HF | 12.20, -11.71, -8.05 | BSC 2202 |
| S27 | L | 6X | M | LF and HF | -10.78, -13.67, -11.39 | MDT B33005 |
|  | R |  |  | LF and HF | 12.40, -13.54, -9.34 | MDT B33005 |
| S28 | L | 6X | F | LF and HF | -13.00, -11.60, -7.63 | BSC 2202 |
|  | R |  |  | LF and HF | 13.34, -11.89, -8.63 | BSC 2202 |
| S29 | L | 6X | M | LF and HF | -13.37, -12.30, -7.82 | MDT B33005 |
| S30 | L | 5X | F | LF and HF | -13.45, -14.50, -7.45 | BSC 2202 |
|  | R |  |  | LF and HF | 12.60, -15.33, -8.28 | BSC 2202 |
| S31 | L | 5X | F | LF and HF | -11.57, -14.11, -8.80 | BSC 2202 |
|  | R |  |  | LF and HF | 10.54, -14.13, -9.28 | BSC 2202 |

Hemi = hemisphere; MNI = Montreal Neurological Institute; MDT= Medtronic; ABT= Abbott; BSC = Boston Scientific

### 2.2. Surgical and recording setup

Quadripolar DBS leads (Medtronic 3389) and directional leads (Abbott Infinity 6172, Medtronic B33005, Boston Scientific Vercise Cartesia Directional Lead 2202) were implanted under standard MER guidance and surgical procedure[6, 32]. As part of the standard functional-mapping MER-based procedure, an expert electrophysiologist determined the depth of STN dorsal and ventral borders and nucleus span within each trajectory where DBS leads were subsequently implanted, blinded to DLEP and spectral measures. The clinical electrophysiologist also annotated the location of each level of the DBS lead based on the final trajectory mapping [33, 34, 35, 36].

The intraoperative local field potential (LFP) data from DBS leads were acquired with the Neuro Omega electrophysiology system (Alpha Omega, Israel) as monopolar recordings from each ring or segment contact, connected through custom adapters. Scalp needles were used for reference and ground.

Recordings were conducted at least 12 hours after discontinuing all anti-parkinsonian medications and at least 30 minutes after terminating propofol sedation, with the patient fully alert and awake, as part of the standard clinical protocol.

While different DBS leads from multiple manufacturers were used, all shared the same contact spacing (1.5mm ring height, 0.5 mm spacing between ring contacts). All the directional leads had standard 1-3-3-1 configuration (4 levels), where the two levels in the middle of the lead are split in 3 directional contacts. We considered all directional and non-directional leads as quadripolar (1-1-1-1 configuration), averaging signal from directional contacts on the same level, as detailed in Signal Analysis, and therefore not considering directional information for the scope of this work.

Additionally, we recorded a subset of our data (5 patients, 9 sides) with a parallel system able to record from contacts under stimulation (Tucker-Davis Technologies, TDT, Alachua, FL, USA), while using NeuroOmega system solely for stimulation. This was possible through a Y-splitter and the higher dynamic range of the TDT recording system (±500mV), and the stimulator having a separate grounding (shoulder) compared to the recording (scalp). This additional data was used solely to validate the DLEP recovery model described in the “DLEP estimation at the stimulating contact” section.

### 2.3. Experimental stimulation paradigm

Electrical stimulation was delivered intraoperatively through the implanted DBS leads using the NeuroOmega clinical system, connected through custom adapters. Stimulation was delivered in a monopolar configuration using a 2×4 inch surface electrode on the shoulder as the stimulation return. The recording protocol consisted of 3 phases: baseline at rest with no stimulation, stimulation with individual pulses delivered at low frequency, stimulation with high-frequency bursts (subset of 25 hemispheres, as specified in Table 1). Specifically, the second phase consisted in delivering individual pulses at low-frequency (LF-DLEP) at 10Hz for 12 s with 3s pause between stimulation settings, through each lead level (ring or pseudoring) in randomized order (Fig. 1B top, Suppl. Fig. 1A). The third phase consisted of delivering burst stimulation, where each burst was composed of 11 stimulation pulses at 130Hz, with 100ms interval between each burst and repeated 30 times for each level, with 3 seconds pause while cycling through each lead level in randomized order (Fig. 1B bottom, Suppl. Fig. 1A). In all protocols stimulation was delivered with an asymmetric biphasic waveform at 1mA, 60µs pulse width for the first phase, 70µs interphase duration, and 480µs for the second phase, to approximate clinical pulse generators.

### 2.4. Signal Analysis

All analyses were performed offline using Matlab version 2023a software (Mathworks Inc., Natick, MA, USA), including its statistical package. All data used for power spectral density (PSD) computation was visually inspected for artifacts or electrical noise. No channels were excluded from analysis, only excluding portion of the signal if it presented unrecoverable artifacts. At least 60 seconds of data was used for each PSD computation.

Electrophysiology was only considered at each ring/pseudo ring level, and we did not consider directional contacts individually. In the case of directional contacts, we averaged signal across all directional contacts within the same level (defined as a pseudoring).

### 2.4.1. DLEP signal analysis and artifact removal

To maximize the spatial sampling through stimulation evoked activity (DLEP), we aimed to recover DLEP from a monopolar montage, during each stimulation block from non-stimulated channels. Importantly, the signal did not saturate in the non-stimulated channels after 1ms of each stimulation pulse, which allowed stimulation artifact removal. To estimate and remove the stimulation artifact due to electrode-tissue interface[37], we expanded upon the assumption that the tissue-electrode interface acts as a 1^st^ order filter (RC filter) [38].

Specifically, we assumed an electrical circuit equivalent (Suppl. Fig. 2), based on prior work on concurrent stimulation and recording [39], keeping only the circuit path for the monopolar case (one electrode stimulating and one electrode recording). Due to its configuration, the equivalent circuit can be seen as a canonical 2^nd^ order system, so in turn its step response can be expressed in the time domain as a damped sinusoid[40, 41]. As stimulation pulse can be seen as a summation of step responses[42], it will in turn give rise to a summation of damped sinusoidal responses. As the largest contribution is from the first cathodic phase, we used a maximum of 3 damped sinusoids for fitting the artifact, defined as: 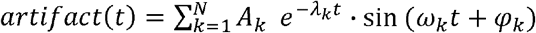, where *A*_*k*_ is the magnitude of the dampened sinusoid, *λ*_*k*_ is the decay of the exponential component, *ω*_*k*_ and *φ* _*k*_ are respectively the frequency and phase shift of the sinusoidal component.

Furthermore, to ensure we did not alter the physiology of the evoked potential of interest (DLEP with its oscillatory frequency between 250-530Hz)[20, 22, 43], we constrained the fitting to contributions with a frequency content below <160Hz. Thus, we have estimated and removed the stimulation artifact by fitting a family of constrained dampened sinusoids to the artifact, using a nonlinear optimization with the goal of minimizing the mean squared error (MSE) between the original noisy data and the fitted artifact. We leveraged the nonlinear fit function “lsqnonlin” from Matlab. To smoothly remove slow baseline drift we leveraged a Savitsky-Golay filter at a wide window (6.7ms)[30, 44]. Thus, this method allows separating the electrophysiology of interest from the artifact induced by stimulation (Fig. 1D).

Once the artifact was removed from the averaged evoked potential, the DLEP amplitude was measured as the root-mean-square of the envelope of the signal (4-20ms after stimulation pulse for low-frequency stimulation or after burst end for high-frequency stimulation), for each channel/contact that was not under stimulation. Data was not recorded from the contact being stimulated, aside from a subset of 5 subjects with an additional recording system (TDT).

The DLEP oscillatory frequency for each stimulated channel was measured on the non-stimulated contact with the maximum DLEP amplitude, as the inverse of the average time interval between the first two consecutive DLEP peaks. To ensure a stable DLEP oscillatory frequency measure, we included contacts with DLEP amplitude of at least 4µV RMS, discarding ~17.11% of the available contact data. All values for DLEP amplitudes were z-normalized within each patient and side for further group analysis, aside from when otherwise specified (e.g. 2D/3D spatial plots).

#### 2.4.2. DLEP estimation at the stimulating contact

As most recording systems including NeuroOmega cannot record from the same contact that is being stimulated, we aimed to accurately predict the DLEP amplitude from the contact delivering stimulation. Based on our initial observations, we postulated that DLEP amplitudes would follow a normal distribution centered on the same STN location regardless of the contact being stimulated.

Hence, to account for variability of the sampled DLEP amplitudes across multiple stimulations, we leveraged a generalized normal distribution, specifically the skew normal distribution[45, 46]. As shown in previous work, local field potentials can spread across neighboring regions, showing a decay of LFP amplitudes following a gaussian spread[47, 48]. The skew normal distribution, also known as skewed gaussian, assumes a random variable *X* follows the probability density function (pdf): *f*_*SN*_ (*x*) = 2*ω*^-1^ φ (*z*) Φ (*α z*) where *z*= *ω*^-l^(*x* - *ξ*), *φ* is the *N* (0,1) density (pdf of normal distribution), Φ is the cumulative distribution function of *N*(0,1), and *α* its skewness parameter [49, 50]. Specifically, its parameters can be defined in function of its own mean (μ) and standard deviation (*σ*), leading to location (*ξ*) as 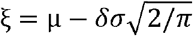 and scale *ω* as 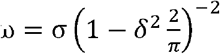, where 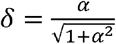 [51].

Considering that each lead was subdivided in 4 levels (1-1-1-1 configuration), for each level that was stimulated we collected DLEP activity at the 3 adjacent levels. Thus, we obtained 3 amplitude measures for each stimulated level, for a total of 12 DLEP amplitudes.

We then fitted one skewed gaussian for every 3 amplitudes from each stimulated level, for a total of 4 skewed gaussians. To ensure a unique fit, we linked the 4 fitted skewed gaussians by constraining to have the same mean (μ) and variance (*σ*), allowing the skewness parameter (α) to be independent for each curve. As consequence, each stimulation level had one curve fitted with their own unique location (*ξ*) and scale (ω).

To find the best fitting skewed gaussians across the 4 sampled levels, we used a nonlinear optimization with the goal of minimizing the MSE between the original sampled amplitudes and the amplitudes obtained from each curve fit. We leveraged the nonlinear fit function “lsqnonlin” from Matlab. For the fitted values we considered only the locations sampled in the original data (3 amplitudes for each stimulated level). Once obtained the optimal family of 4 skewed gaussians with shared μ and *σ*, we interpolated the missing DLEP amplitudes on the stimulating contact. Detailed workflow is described in Supplementary Figure 1.

To validate our model, we recorded a subset of our data (5 patients, 9 hemispheres) with a parallel system able to record from contacts under stimulation (Tucker-Davis Technologies, TDT, Alachua, FL, USA). Neuro Omega system was still used to deliver stimulation and record from non-stimulating contacts. We compared the model-predicted DLEP amplitude with the one that was directly recorded by TDT, for both low-frequency stimulation (Fig. 2B, Suppl. Fig. 3A), and high-frequency stimulation (Fig. 2C, Suppl. Fig. 3B). We show that there is a high correlation between predicted and recorded DLEP amplitudes (on stimulated contacts), for both low-frequency stimulation (Fig. 2B, R^2^adjusted=0.834, p-val<0.001), and for high-frequency stimulation (Fig. 2C, R^2^adjusted=0.920, p-val<0.001).This demonstrates that it is possible to accurately estimate DLEP activity using a common clinical amplifier, even when not able to record from the same contact that is being stimulated, and only having information from adjacent contacts. The DLEP amplitudes at the stimulating contacts in the remainder of this work were obtained from this model. DLEP hotspot was defined as the location with the highest DLEP amplitude for each hemisphere.

**Figure 2.**
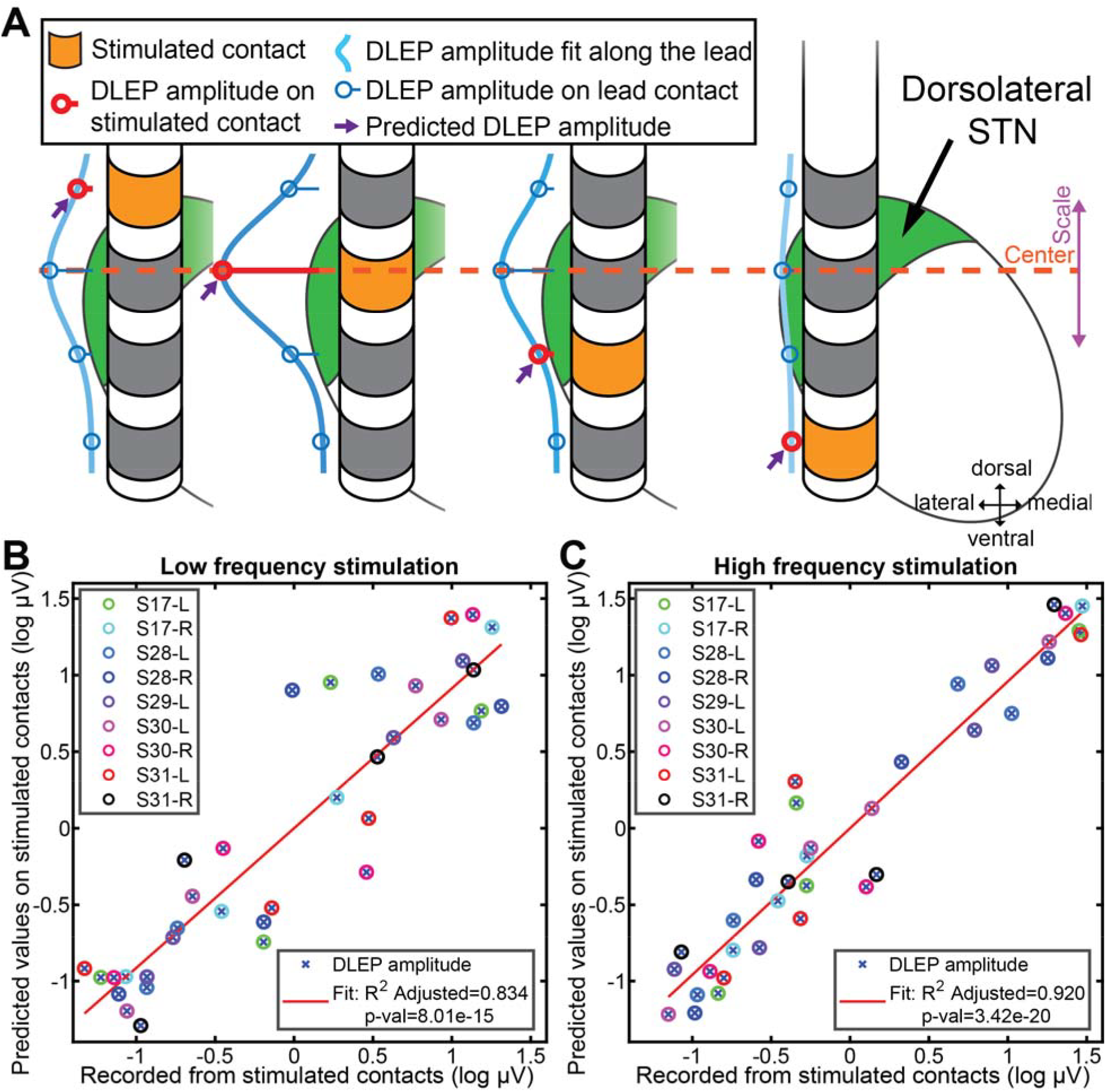
DLEP relationship with STN and DLEP recovery. (A) Distribution of STN-DLEP amplitudes depends on stimulating contact (orange). DLEP hotspot is defined as the location (i.e. contact) where stimulation elicits the highest DLEP amplitude. The highest amplitude is hypothesized to be recovered at the contact within dorsal-STN (as shown in this example). A mathematical model is used to fit gaussian curves to the distribution of DLEP responses and estimate the DLEP amplitude at the stimulated contact (light blue line). This method is used to predict the amplitude of DLEPs from channels not recorded due to stimulation (highlighted in red), estimating the amplitude at those contacts (violet arrow). We validated the model-based fitting through additional hardware (TDT) capable of recording from the stimulated contacts in a subset of patients. With the clinical system (Neuro Omega), the channel stimulated is not recoverable due to saturation. Across 5 patients tested (9 hemispheres), the recovery of the DLEP at the stimulated contact for both (B) low frequency stimulation and (C) burst of high frequency stimulation had a strong and significant correlation, indicating the validity of the mathematical model (LF R^2^=0.834, p-val<0.0001, HF R^2^=0.920, p-val<0.0001).

#### 2.4.3. Spectral analysis

Using resting data when no stimulation was delivered, we calculated the power spectral density (PSD) for each monopolar contact using a Fast Fourier Transform (FFT) with the Welch method (1 s Hamming window, 80% overlap). We used the irregular-resampling auto-spectral analysis (IRASA)[52] method to parametrize the spectra, separating the aperiodic from the oscillatory component. We then automatically determined the peak within the beta range (13-35Hz) with “findpeaks” function from Matlab. A peak was always found within the beta range, but peak prominence was not considered in our analysis. From the found beta-peak frequency, we computed the average beta peak power considering a bandwidth of 5Hz. We also computed the high frequency oscillation (HFO) power in the 250-350Hz band, centered on the salient 300Hz fast oscillation[53].

The IRASA method allowed us to take into account patient variability, and oscillatory power values were normalized based on each channel aperiodic amplitude offset. Since we extracted the aperiodic component of the spectra, we also considered the aperiodic power between 8-350Hz as an additional spectral measure for subsequent analysis (lower limit for alpha rhythm at 8Hz to upper limit for HFO at 350Hz). Due to line noise (60Hz and harmonics) and narrowband artifacts given by the intraoperative O-Arm (Medtronic, USA) (105.5Hz and harmonics), we removed spectral bins at the respective aforementioned frequencies for calculating HFO power (5Hz bandwidth). The aperiodic estimate is not affected by these narrowband artifacts by construction [52].

All values for spectral amplitudes (beta, HFO, aperiodic) were z-normalized within each hemisphere for further group analysis, aside from when otherwise specified (e.g. not-z-normalized 2D/3D spatial plots). When “not-z-normalized” is indicated, this means beta and HFO power were relative to the aperiodic power (i.e. normalized to just aperiodic power), while aperiodic component of the spectra was the absolute power.

### 2.5. Lead locations and trajectory localization

Lead locations were obtained both from clinical intraoperative functional-mapping (MER-based) and from post-operative imaging. For comparison across patients we normalized the MER-based span of the STN, defining as 0 the ventral border, and 1 as the dorsal border. We grouped locations in bins with increments of 0.25. Locations above 1 are locations above the dorsal border.

For patient imaging, lead locations were obtained using the LeadDBS processing pipeline (v3) [54, 55] with the following steps. Pre-operative MRI (T1, T2, fGATIR if available, with T1 as primary reference image) and post-operative CT were linearly coregistered using advanced normalization tools (ANTS) [56], applying a brain-shift correction [57], and inspected through LeadDBS pipeline. To compare across patients, we normalized each patient’s brain to a normative Montreal Neurological Institute (MNI) space (ICBM 152 Nonlinear 2009)[58, 59] through Lead-DBS toolbox[55], using ANTs SyN Diffeomorphic Mapping [60] with pre-validated presets[61]. DBS leads were automatically pre-reconstructed and localized using PaCER method[62] and manually refined through visual inspection, if needed.

We extracted each lead contact imaging-based coordinate directly within MNI space. We then centered all lead contacts coordinates with respect to STN center, which is considered as the origin (0,0,0) coordinate. The locations were exported to MATLAB for further analysis together with electrophysiological measures (e.g. DLEP amplitude, spectral power). Coordinates for the leads implanted in the left hemisphere were mirrored in the right hemisphere to allow for comparison regardless of hemisphere recorded (Suppl. Fig. 4A). Lead trajectories were identified based on contact locations within each lead.

For the purpose of comparing electrophysiological measures (e.g. DLEP amplitude, spectral power) across patients only as a function of depth and STN borders, we obtained the average trajectory across all the lead contacts, and projected each electrophysiological measure on the average trajectory itself (to a location orthogonal to the average trajectory from each identified contact) (Suppl. Fig. 4B). Thus, we obtained a 1D mapping of the electrophysiological measures, together with STN borders locations. The borders of the STN were extracted from the DISTAL atlas[63, 64] coregistered to the MNI space. The STN borders are localized as the intersection between the STN boundaries from the DISTAL atlas[63, 64] and the average lead trajectory. Thus, the ventral border and dorsal border were localized on the 1D mapped locations at −2.9003mm and at 3.9115mm, respectively. The projected center of the STN was set at 0mm. Data were clustered in 2mm bins centered on the clustering depth (e.g. −1mm).

For 3D interpolation across locations/electrophysiological measures, we used a scattered linear interpolant (MATLAB scatteredInterpolant() with Natural neighbor interpolation), only within a volume region defined by the sampled locations (Suppl. Fig. 4A), therefore no extrapolation was considered. Points were interpolated within a fine equidistant grid with 0.1mm spacing. To better visualize a distribution of electrophysiological measures across contacts, with respect to depth for the purpose of DBS targeting, we used the average trajectory to find the plane orthogonal to the axial plane and containing the average trajectory line itself (Suppl. Fig. 4B), defined as the trajectory plane.

### 2.6. Clinical outcomes measures

We evaluated clinical efficacy of each lead contact by performing retrospective chart review. Specifically, one investigator (SM) reviewed clinical programming notes and scored each of the four lead rings/pseudorings based on clinical effectiveness described by the programming clinician during monopolar mapping so that 0=no effect and 4=maximum benefit (e.g. a contact with clinical effectiveness at a lower stimulation amplitude was scored higher than a contact with similar clinical effectiveness at a higher amplitude). Interpreting investigator and clinical programmers were not aware of electrophysiologic study findings and LFP sensing was not used in clinical programming. If a contact was not evaluated, it was not assigned a value and was excluded from clinical outcome analysis. A few leads did not have adequate clinical notes to allow contact ranking (4 of 39 leads for LF-DLEP, 2 of 22 leads for HF-DLEP). To compare across patients, we scaled the within-patient ranking, mapping it from 0 (no effect) to 1 (maximum clinical efficacy) across contacts within the same lead. The normalized values were binned (0-0.25, 0.25-0.5, 0.-5-0.75, 0.75-1) and each bin assigned a rank (worst clinical efficacy as rank 4 to best clinical efficacy as rank 1).

### 2.7. Statistical analysis

Electrophysiologic measures (DLEP amplitude, spectral power) were log-scaled to compensate for the power-law right-skewed distribution[52]. Unless specified, data (DLEP amplitude, spectral power) was z-normalized within each lead across patients. A p-value <0.05 was considered statistically significant. All correlations are Pearson and all p-values are two-tailed, unless indicated otherwise as for clinical efficacy (Spearman). In the case multiple pairwise correlations were computed across estimates (spectral measures vs DLEP, Figure 4) within the same context, correlation significance was adjusted with Bonferroni[65]. Non-parametric Wilcoxon rank-sum test was used in pairwise comparison tests without assumption for normality (for clinical efficacy), adjusted for multiple comparisons with Holm-Bonferroni method[66]. Comparisons of DLEP and spectral amplitudes across locations were performed with non-parametric ANOVA on ranks (Kruskal-Wallis test) as it does not assume normality and we considered the binned locations as sampled independently, similarly to previous studies[12, 23, 67, 68, 69]. Post-hoc analyses were corrected for multiple comparison through Dunn-Sidak method.

## 3. Results

We recorded local field potentials (LFP), obtaining DLEP and spontaneous spectral measures (beta, HFO, aperiodic component of the spectra) in 31 subjects affected by Parkinson’s Disease (25 men, 6 women; average age at surgery 57.8±8.5 years) who underwent MER-guided STN DBS implantation, resulting in a total of 39 hemispheres recorded. Clinical characteristics and experimental setup are detailed in Table 1.

We used a novel signal processing algorithm to evaluate DLEP at each DBS contact including estimating DLEP at the stimulating contact (Fig. 2). This allowed a high-resolution analysis of DLEP spatial distribution across the STN, facilitating the comparison with spectral measures to evaluate the utility for targeting and postoperative programming.

### 3.1. Stimulation frequency influenced DLEP morphology but not spatial distribution

Regardless of stimulation frequency (low-frequency pulses vs high-frequency bursts), the distribution of DLEP amplitudes across each patient’s lead showed high correlation (Fig. 3A, R^2^-adjusted=0.632, p-val<0.0001), supporting that the spatial mapping across contacts is preserved with both low-frequency and high-frequency stimulation. This effect was even stronger when normalizing data points within each lead (z-normalization), removing the effects of overall DLEP amplitude differences across patients/hemispheres (Fig. 3B, R^2^-adjusted=0.751, p-val<0.0001). This relationship was maintained despite the significantly larger amplitude (LF=20.92 (SD=14.82)µV, HF=53.57 (SD=39.96)µV, p-val<0.0001) for high-frequency burst stimulation compared to with individual pulses delivered at low frequency. Importantly, even with LF evoked DLEP, which in past work have been defined as “non-resonant” [22, 30, 31], the spatial information is retained.

**Figure 3.**
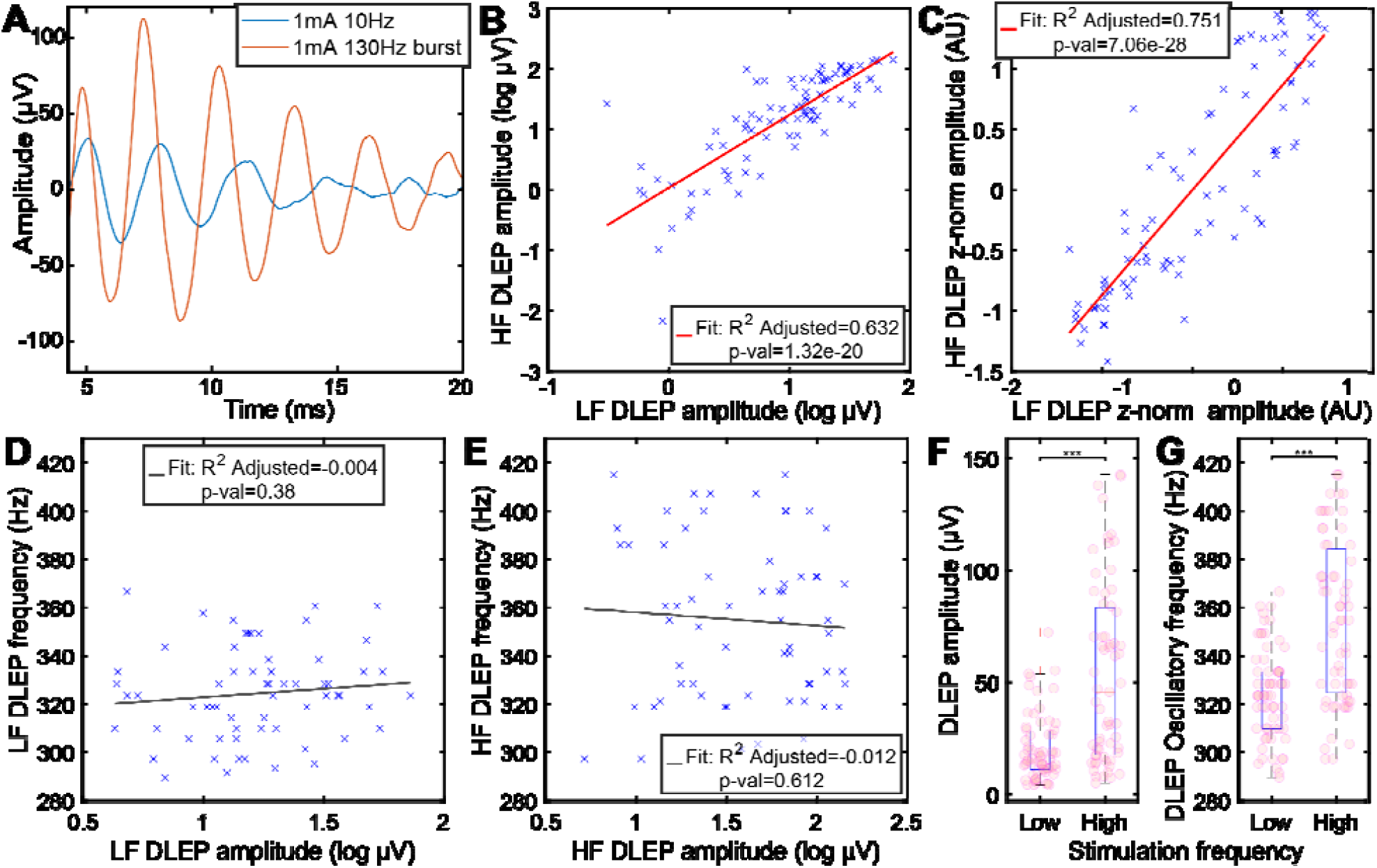
Comparison of DLEP morphology elicited with low frequency and burst of high frequency stimulation. A) DLEP amplitudes (not z-normalized) showed high correlation across stimulated contacts (R^2^-adjusted=0.632, p-val<0.0001) between low (10Hz) and high frequency burst stimulation (130Hz). The not-z-normalized (DLEP absolute amplitude) data is used for spatial plots across locations, and here is shown for reference. B) Normalized DLEP amplitudes showed high correlation across the stimulated contacts (R^2^-adjusted=0.751, p-val<0.0001), highlighting that the DLEP amplitude trend across contacts within each lead was maintained in the two stimulating conditions, ignoring effects of overall DLEP amplitude differences across patients/hemispheres which does not provide further depth information. C) Example differences between DLEP morphology in a single patient between low and high frequency stimulation. D)E) There was a lack of correlation between DLEP amplitude and DLEP oscillatory frequency at both low frequency (D) and high frequency (E). F) The DLEP amplitude is significantly larger (208.58%, SD=324.41%, p-val<0.0001) for high frequency stimulation compared to low frequency stimulation. G) The DLEP oscillatory frequency is significantly different depending on the frequency of stimulation used (p-val<0.0001). Higher frequency of stimulation evokes DLEP oscillatory frequency (9.66%, SD=11.42). In all the analysis (D,E,G) with DLEP oscillatory frequency we used a subset of patients/leads that had both LF and HF DLEPs (48.72%), and with a minimum of 4µV RMS to allow for a stable frequency estimate (82.89% of the paired contacts, 40.38% of the total contacts).

**Figure 4.**
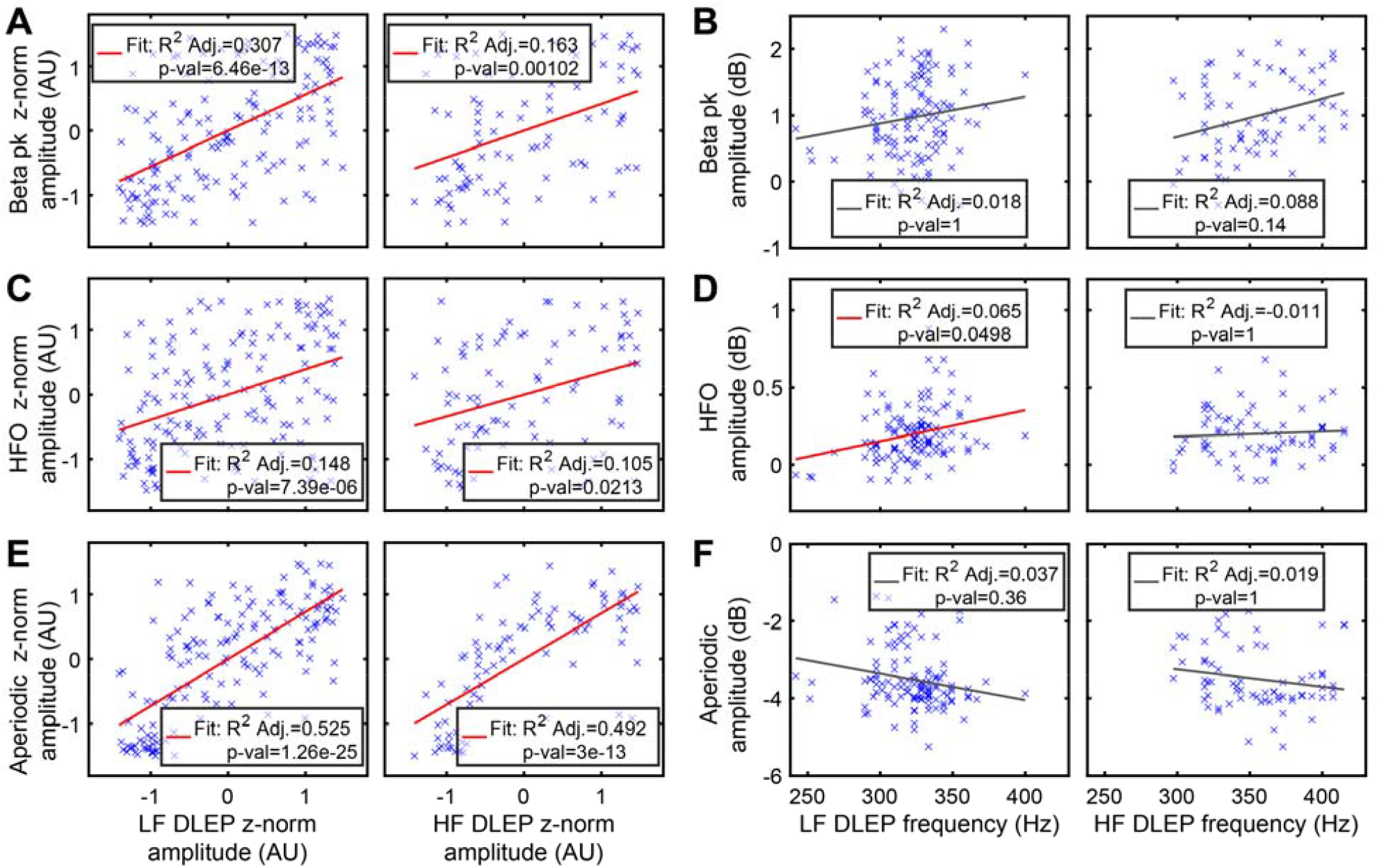
Relationship between DLEP morphology and spectral features of resting-state LFP signal. We compared DLEP amplitude and DLEP oscillatory frequency with spectral components obtained from resting-state (no stimulation) LFP signals: (A,B) beta, (C,D) high frequency oscillations (HFO), (E,F) high frequency aperiodic component. Left column is LF evoked DLEP, right column is HF evoked DLEP, for each subpanel. In A,C,E DLEP amplitudes are normalized across patient and leads to highlight relationships between amplitude trend across contacts. In B,D,F we compare overall spectral feature strength with DLEP oscillatory frequency. While there is a significant weak correlation between beta (A, LF DLEP R-Adjusted^2^=0.307, p-val<0.001; HF DLEP R-Adjusted^2^=0.163, p-val<0.01), and HFO (C, LF DLEP R-Adjusted^2^=0.148, p-val<0.001; HF DLEP R-Adjusted^2^=0.105, p-val<0.05) spectral amplitude and DLEP amplitude, there is a higher correlation (E) between DLEP amplitude and aperiodic component (LF DLEP R-Adjusted^2^=0.525, p-val<0.001; HF DLEP R-Adjusted^2^=0.492, p-val<0.001). There was a significant trend between HFO amplitude and oscillatory frequency, which however, had a negligible effect explained (D left, LF DLEP R-Adjusted^2^=0.065, p-val<0.05). No significant trend was found between LF DLEP oscillatory frequency and both beta and aperiodic (B,F left). For HF DLEPs instead there was no significant relationship with all spectral estimates beta, HFO, and aperiodic (B,D,F right). In all the analysis (D,E,G) with DLEP oscillatory frequency we used a subset of contacts that had DLEPs with a minimum amplitude of 4µV RMS to allow for a stable frequency estimate (82.89% for LF and 80.23% for HF DLEP contacts).

At the same time, stimulation frequency influenced the oscillatory frequency of the elicited DLEP (Fig. 3C,3F), with higher stimulation frequency increasing (LF=324.41 (SD=19.16)Hz, HF=354.81 (SD=33.38)Hz, p-val<0.0001) the DLEP oscillatory frequency (Fig 3F). This might be a consequence of a compounding effect occurring with high-frequency stimulation[24]. However, the DLEP oscillatory frequency was not related to the strength of the evoked response (DLEP amplitude) regardless of stimulation frequency (Fig 3D,3E). This suggests that DLEP frequency is independent of DLEP amplitude.

### 3.2. Spontaneous oscillatory activity distribution suggests a weak overlap with DLEP generators

We evaluated spontaneous oscillatory activity while the patient was resting and no stimulation was delivered. We focused on beta peak (13-35Hz) and HFO (250Hz-350Hz). Analysis showed that z-normalized oscillatory beta peak (Fig. 4A) amplitudes moderately correlate with z-normalized DLEP amplitudes, regardless of type of stimulation evoking it (LF-DLEP R-Adjusted^2^=0.307, p-val<0.001; HF-DLEP R-Adjusted^2^=0.163, p-val<0.01). Similarly, beta peak amplitude (not normalized) did not show a strong correlation with DLEP amplitude for both paradigms (Suppl. Fig. 5A). Furthermore, spontaneous beta peak activity did not show a significant relationship between its amplitude and DLEP oscillatory frequency, regardless of stimulation frequency (Fig. 4B). Similarly to beta, HFO activity showed a significant but weak correlation with DLEP (Fig. 4C), regardless of stimulation type (LF-DLEP R-Adjusted^2^=0.148, p-val<0.001; HF-DLEP R-Adjusted^2^=0.105, p-val<0.05). However, this relationship was maintained for only LF-DLEP for absolute (not normalized) amplitudes (Suppl. Fig. 5B, LF-DLEP R-Adjusted^2^=0.153, p-val<0.001; HF-DLEP R-Adjusted^2^=0.075, p-val=0.102). A very weak relationship was present between HFO amplitude and DLEP frequency (low-frequency stimulation, LF-DLEP Fig. 4D left, R-Adjusted^2^=0.065, p-val<0.05), while absent for high-frequency stimulation (burst, HF-DLEP Fig. 4D right, R-Adjusted^2^=0.011, p-val=1). Overall, the weak correlations suggest that DLEP are not necessarily manifesting from the same subregion of the STN that generates beta activity.

### 3.3. DLEP correlates with spontaneous aperiodic component of the spectra

We evaluated spontaneous aperiodic component of the spectra while the patient was resting and no stimulation was delivered. Aperiodic component has been suggested as a proxy of STN neuronal excitability [70, 71, 72, 73], and it is estimated as shift in broadband power (between 8-350Hz) of spectral fractal component using the IRASA method[52]. We found that aperiodic component amplitude had a significant relationship (Fig. 4E) with z-normalized DLEP amplitude regardless of stimulation paradigm evoking it (LF-DLEP R-Adjusted^2^=0.525, p-val<0.001; HF-DLEP R-Adjusted^2^=0.492, p-val<0.001). This relationship was maintained when considering absolute amplitudes (Suppl. Fig. 5C), showing however that the amplitude relationship is weakly but necessarily related to the overall strength of the aperiodic rhythm/DLEP (LF-DLEP R-Adjusted^2^=0.095, p-val<0.001; HF-DLEP R-Adjusted^2^=0.044, p-val=0.0272). Compounded with the z-normalized findings, this indicates that DLEP and aperiodic generators may overlap, suggesting that DLEP relies on the availability of entrainable STN units, and supporting prior-modeling based findings[20]. Furthermore, the aperiodic component did not show a salient relationship with the overall DLEP frequency (Fig. 4F left, R-Adjusted^2^=0.037, p-val=0.36; Fig. 4F right, HF-DLEP R-Adjusted^2^=0.019, p-val=0.13).

### 3.4. DLEP shows better agreement with MER-based functional-mapping than spectral features

Standard functional-mapping performed by an expert neurophysiologist takes in consideration the underlying pattern variation of single unit activity to define the borders and span of the STN[33, 34, 35, 36]. We compared the performance of DLEP signal and salient spectral power estimates (beta peak, HFO, aperiodic component) in discriminating along the STN borders defined by MER (Fig. 5A).

**Figure 5.**
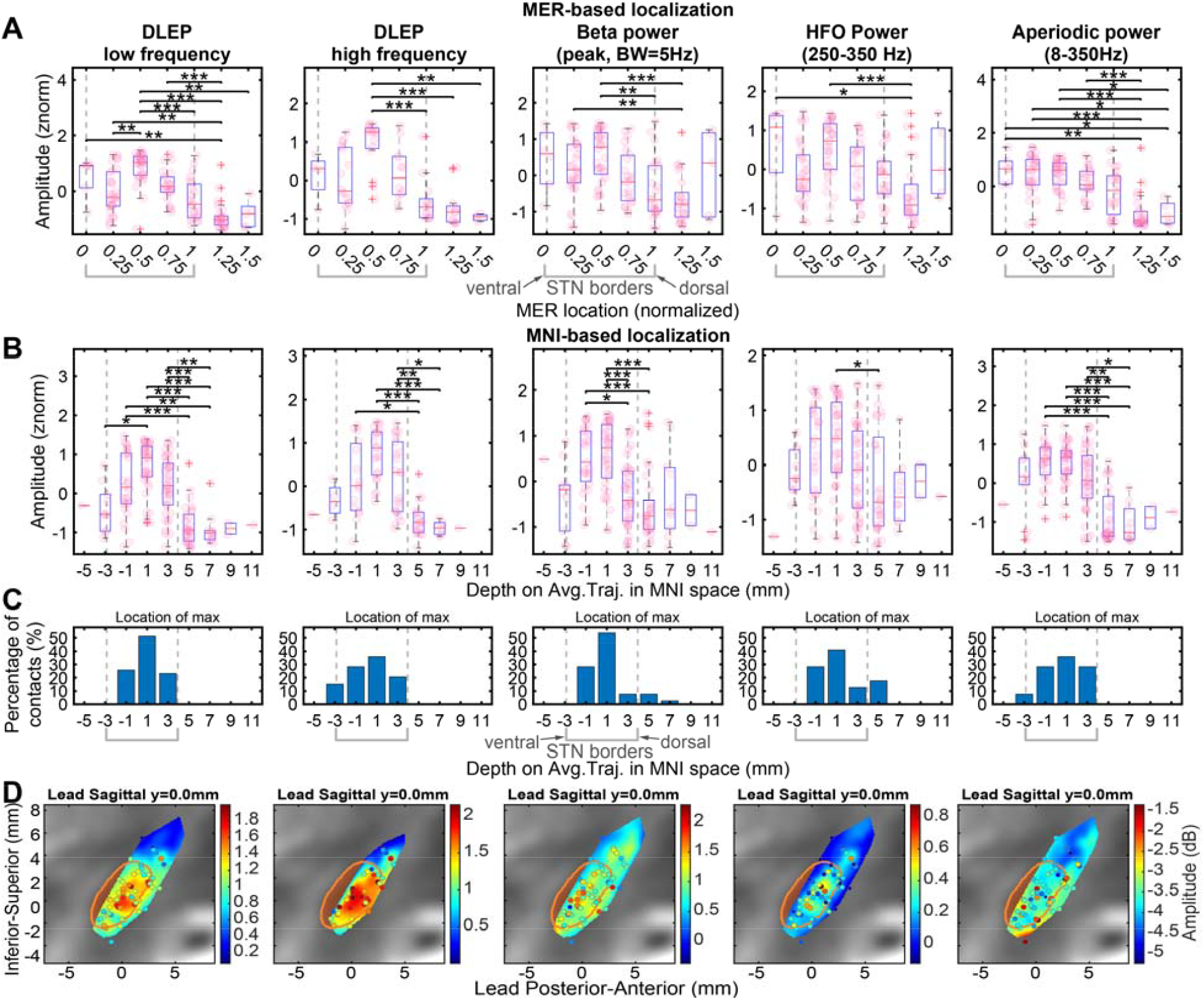
Spatial distribution of DLEP and spectral features. Each graph column represents one estimated electrophysiological marker, in order: LF DLEP, HF DLEP, Beta power, HFO power, power of the aperiodic component of the spectra. Each row shows the distribution of the estimated amplitudes across the electrophysiological makers linked to a different spatial estimate. Row (A) leverages MER mapping to define spatial locations. MER mapping location is normalized across patients and leads between ventral (block 0) and dorsal (block 1) border. Row (B) and (C) leverage normalization to a MNI space and normative atlas. Contact locations are projected to the average trajectory across all the leads, and STN borders are localized by the intersection between the STN atlas structure with the average lead trajectory, at locations −2.9003mm for ventral border and 3.9115mm for dorsal border. (B) shows the distribution of amplitudes across the MNI-based projected location. Row (A) and (B) indicate significant post-hoc pairwise comparisons with a star (1 star p-val<0.05, 2 stars p-val<0.01, 3 stars p-val<0.001).(C) shows the location of the maximum amplitude for each lead of the considered electrophysiological marker. (D) shows the distribution of amplitudes mapped within the MNI space. The MNI space is rotated to have the sagittal plane parallel and including the average lead trajectory (preserving the z-axis, Suppl Fig. 3B). The STN structure contour is shown overlayed to a reference T2 MNI image. The amplitudes of each electrophysiological markers are interpolated with a scattered interpolant. Sampled contacts are shown within ±0.8mm from the shown slice.

Both LF and HF-DLEP had higher normalized amplitudes within the STN border compared to outside of it (non-parametric ANOVA, p-val<0.001). LF-DLEP had the highest number of normalized spatial bins along the MER-mapped STN with significant discrimination, compared with all the other estimates. This was most notably demonstrated at the highest median DLEP amplitude (within STN bin at 0.5, Fig. 5A 1^st^ quadrant), showing a significant difference with areas dorsal to the STN border (p-val<0.001), and ventral areas at bin 0.25 (p-val<0.01). This supports that DLEP are stronger within the dorsal portion of the STN, and decay in amplitude especially outside of the STN. Similarly, HF-DLEP also had highest median amplitude within STN at bin 0.5 (Fig. 5A 2^nd^ quadrant), showing significant discrimination with areas dorsal to the STN border (p-val<0.001), but not at the ventral border.

Spectral features (beta, HFO, aperiodic component) also showed differences across contacts (non-parametric ANOVA, p-val<0.001). Spontaneous beta activity (normalized beta power) showed the highest median within STN at bin 0.5, similar to the DLEP estimates (Fig. 5A 3^rd^ quadrant). This location showed significant discrimination with part of the dorsal STN border (bins at 1, 1.25 with p-val<0.01), but not at the ventral border.

Normalized HFO power also had the highest median within the STN at bin 0.5, but that location showed significant discrimination only with a small portion of the area dorsal to the STN (bin at 1.25, p-val<0.001). Interestingly, the amplitude of the normalized aperiodic component of the spectra did show discrimination across all bins within the STN and the area dorsal to the STN border (p-val<0.05).

### 3.5. DLEP has a higher selectivity in spatial localization along trajectories compared to spectral estimates

Accurate localization within target structures for surgical implantation is a key determinant for optimal therapeutic outcomes, such as sensorimotor region in the dorsolateral STN for PD. Similarly, as for MER-based functional-mapping, we compared DLEP and spectral estimates with imaging based localization. To compare across patients in relation to depth (Fig. 5B,5C), we considered the projected contact location to the average trajectory across all leads. Both LF and HF-DLEP showed higher normalized amplitudes within the STN border (non-parametric ANOVA, p-val<0.001). LF-DLEP amplitudes had the highest discriminability across depths and showed the highest median amplitude towards the dorsal locations (1 and 3mm) (Fig. 5B). DLEP within the STN (−1 to +3mm) were significantly different from areas dorsal to the STN border at 5-7mm (p-val<0.01), and ventral areas at −3mm (p-val<0.05). Similarly to MER-mapped locations, this supports that DLEP has the largest amplitude within the STN. Similarly, HF-DLEP also showed the dorsal STN locations with the highest median (1 and 3mm), but showed a significant difference only with areas dorsal to the STN border (5-7mm, p-val<0.01).

Spectral features (beta, HFO, aperiodic component) showed differences across contacts as well (non-parametric ANOVA, p-val<0.001). Spontaneous beta activity (normalized beta power) showed lower discriminability (compared to LF-DLEP amplitude), as the location with the highest median beta power (1 mm) only showed discrimination with dorsal areas (including outside of the STN at 5mm, and within at 3mm, p-val<0.05). Normalized HFO power showed a significant difference only between the location with the highest median HFO power and area dorsal to the STN border (5mm, p-val<0.05). The aperiodic component of the spectra showed discriminability between areas within the center of the STN (−1 to 1mm) and areas dorsal to the STN border (5-7mm, p-val<0.01, with only 3mm-7mm p-val<0.05).

Importantly, as the goal is to localize the best placed contact for each patient, we identified the contact with maximum value for each estimate of interest for all leads. The contact with the highest DLEP is typically within the STN (100% of contacts for LF-DLEP and 84.6% for HF-DLEP) and lower for other estimates (beta: 89.74%, HFO: 82.05%, aperiodic component: 92.31%) (Fig. 5C). The discriminative power of DLEP compared to spectral components is also shown by inspecting the interpolated amplitude values (absolute DLEP amplitude or spectral power, not z-normalized) across the plane sagittal to the DBS lead trajectory (z-axis kept as invariant), having DLEP concentrating within the STN (Fig 5E). Individual values before interpolation (Suppl. Fig. 6) show DLEP amplitudes focusing within the STN, while beta is qualitatively spread across a larger space, matching what is shown quantitatively in Fig. 5C.

### 3.6. DLEP is correlated with clinical efficacy

DLEP amplitudes and power of spectral features (beta, HFP, aperiodic component) were sorted based on clinical efficacy at individual contacts (ranked from worst (4^th^ rank) to best (1^st^ rank), Fig. 6). Clinical efficacy ranking showed significant correlation with LF-DLEP normalized amplitude (Spearman ρ= −0.325, p-val<0.001), while lower for HF-DLEP (Spearman ρ= −0.261, p-val<0.05). All the higher ranked contacts (1^st^, 2^nd^) from LF-DLEP were higher than the worst contacts (3^rd^ and 4^th^, p-val<0.05). For HF-DLEP only rank 1 contacts were significantly different in amplitude than rank 4 contacts (p-val<0.05).

**Figure 6.**
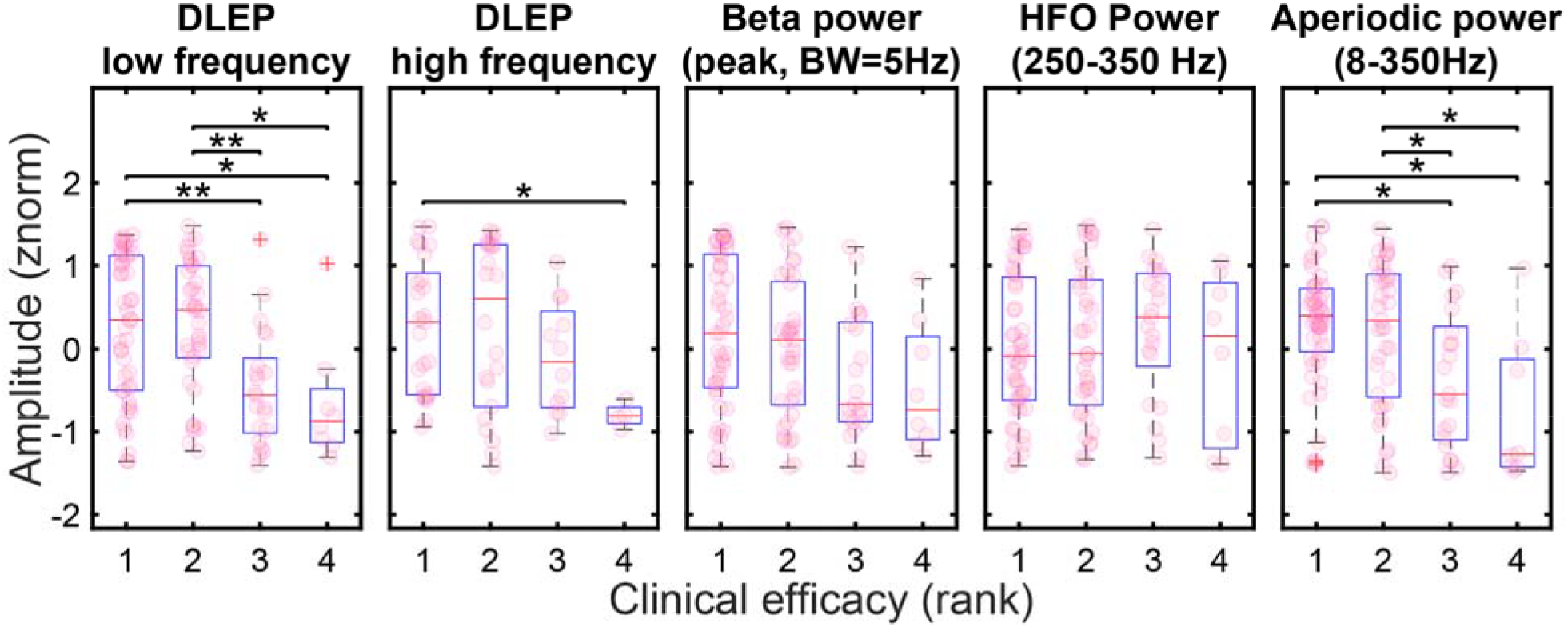
Clinical outcomes across DLEP and spectral features. Contact are ranked by clinical efficacy. DLEP amplitudes and spectral features power are binned within each rank. LF DLEP(ρ= −0.325, p-val<0.001), HF DLEP(ρ= −0.261, p-val<0.05), beta power (ρ= −0.251, p-val<0.01), and aperiodic component (ρ=−0.313, p-val<0.001) power show a significant correlation (Spearman) with clinical efficacy. Significant pairwise comparisons (Wilcoxon rank-sum test, Holm-Bonferroni adjusted) are indicated with a star (1 star p-val<0.05, 2 stars p-val<0.01).

Spontaneous beta activity (normalized beta power) was also significantly correlated (Spearman ρ= −0.251, p-val<0.01), but to a lesser degree than LF-DLEP. It also showed lower discriminability across rankings (no significant difference in beta amplitude across rankings). Spontaneous HFO (normalized) did not show a significant correlation with clinical efficacy (Spearman ρ= 0.051, p-val=0.595). The aperiodic component of the spectra interestingly showed a higher correlation than beta and HFO (Spearman ρ=−0.313, p-val<0.001), showing also significant difference between the highest ranked contacts (1^st^, 2^nd^) ranked contacts and the worst contacts (3^rd^ and 4^th^, p-val<0.05).

## 4. Discussion

This intraoperative study evaluated electrophysiologic characteristics and clinical utility of a relatively novel DLEP marker against well-established LFP oscillatory measures in PD patients. Postoperatively, clinical DBS is applied at high frequencies (>100 Hz) to achieve therapeutic benefit, and most prior studies have investigated DLEP using bursts of high-frequency stimulation.

We first demonstrate that there is no information loss in spatial localization using DLEP elicited with single pulses delivered at low-frequency (LF-DLEP), compared to DLEP elicited with high-frequency bursts (HF-DLEP). Next, we show that DLEP is more accurate in localizing DBS electrode contacts within STN borders than oscillatory measures when using either MER-guided or image-guided clinical mapping as gold-standard. Interestingly, DLEP amplitudes strongly correlate with aperiodic spectral components, suggesting a link between DLEP and underlying neural excitability. Finally, we demonstrate that DLEP is more informative for choosing clinically optimal stimulation contact than other measures.

Importantly, this study introduces and validates novel artifact removal and signal recovery methods to enable DLEP recovery using monopolar montage and standard clinical system, thereby enhancing spatial sampling and enabling clinical translation.

### 4.1. A more comprehensive spatial assessment by recovering DLEP at stimulating contact

To improve spatial specificity of DLEP assessment, we focused on enabling recovery at each contact while using monopolar stimulation. We addressed a few limitations of previous DLEP work [20, 22, 23, 26], such as the use of bipolar-montage, biphasic-symmetric stimulation, re-referencing to contralateral side, research-grade recording amplifiers, and ring-only DBS leads, which were techniques used to limit stimulation artifacts.

At the same time, we also designed and implemented a novel method to estimate the STN DLEP amplitudes at the stimulated contact when recording is possible only in adjacent contacts. This is typically the case in off-the shelf clinical amplifier/stimulators (e.g. Neuro Omega) as used in this study. This approach provides DLEP estimates at each electrode contact to compare with gold-standard postoperative monopolar mapping[74], unlike previous studies that evaluated only middle contacts[20, 26].

### 4.2. DLEP and oscillatory activity are localized but potentially distinct phenomena

Beta activity has shown promise as a targeting marker for surgical STN implantation for PD[3, 75]. Here we show that DLEP (specifically individual pulses delivered at low-frequency) is more accurate for localization than beta power when compared across patients, and not just as group level hotspot[3]. Additionally, comparing DLEP and beta amplitudes sampled at the same locations showed a relatively weak correlation. This may suggest that DLEP and beta activity may arise within slightly different locations due to differences in amplitude distributions across sampled locations. Regardless, they both appear to be qualitatively co-localized within the dorsolateral portion of the STN (Suppl. Fig. 6), even if beta is distributed over a larger area and has higher variability across subjects (Fig. 5), suggesting they may engage related subcortical circuits. On this point, recent modeling studies suggest DLEP arise from reciprocal connections between GPe and STN [20, 28, 31], which has also been suggested as generator for oscillatory beta activity[28, 76, 77, 78, 79, 80, 81, 82]. Other authors have argued that DLEP oscillatory frequency correlates with spontaneous beta activity amplitude and used this as rationale that they are linked phenomena[28, 31]. However, this was solely based on a single experimental study[22], and was not replicated in our study(Fig. 4B).

As DLEP spectral content overlaps with HFOs (250-350Hz), we aimed to dissect the relationship between spontaneous HFO and DLEP. Our results showed a weak correlation between the two, reinforcing recent findings that HFO and DLEP are distinct neurophysiological phenomena [22, 27, 30]. Because DLEP are present in patients with dystonia[83] and healthy non-human primates [84, 85], this suggests that DLEP may be a structural marker of basal-ganglia circuit engagement, and not related to PD pathophysiology [28].

### 4.3. Aperiodic component and excitation/inhibition STN balance

Oscillatory activity, such as beta, has played a central role in describing pathological and behavioral states in PD [13, 14, 15, 16, 17, 53, 82], while aperiodic component of the spectra had been typically relinquished to normalization purposes to allow comparison across patients/contacts. Recent work has drawn attention to aperiodic component of the spectra[52, 70, 86], especially on its physiological significance. During no stimulation or in absence of levodopa, STN neurons may exhibit increased firing due to increased excitatory input from cortex and due to decreased inhibitory input from GPe[70, 81, 87, 88]. Changes in aperiodic spectra in the basal ganglia have been proposed as indicator of the excitatory/inhibitory balance for STN inputs [70, 73], associating its broadband-power increase with increased excitability of STN neurons. This is particularly relevant for DLEP, as they may arise from reciprocal GPe/STN connections [20, 24, 28, 31]. Here we suggest that the relative distribution of aperiodic power within a subject mirrors the neural excitability within STN (and consequently the entrainment of GPe-STN loop), which in turn manifests in larger DLEP amplitudes depending on the location being stimulated.

### 4.4. High-frequency stimulation is not necessary for the purpose of spatial mapping

The majority of DLEP studies have focused on spatial-mapping based on activity evoked through high-frequency stimulation[23, 26, 28, 89]. In this study, we delivered stimulation either as short bursts of high-frequency stimulation, or as low-frequency stimulation where each individual stimulation pulse was considered as a single event evoking a DLEP. We clearly show that individual stimulation pulses delivered at low-frequency are sufficient to generate DLEP activity useable for spatial mapping. Therefore, we argue that high-frequency stimulation is not necessary to induce GPe-STN reciprocal activity, as previously suggested [28].

We found a strong relationship between low-frequency (LF) and high-frequency (HF) DLEP amplitudes, which suggest both arise from similar neural targets. The variance between the two estimates (LF-DLEP and HF-DLEP) could be influenced by a variety of factors, including underlying recording noise and differential sample number. Prior work suggested higher stimulation frequency could present a compounding effect due to interaction between successive pulses[22, 30], and different frequency of stimulation might differentially recruit underlying neural populations according to their underling behavior as intrinsic oscillator[30].

This is also supported by the slightly lower DLEP oscillatory frequency evoked by LF stimulation vs HF stimulation. Regardless, LF-DLEP were the best predictor for STN location (100% accuracy), when compared to any other marker (including HF-DLEP). This is possibly due to smaller sample size for HF-DLEP compared to LF-DLEP. Importantly, single pulse stimulation and recording may be easier to implement in future DBS devices. Single pulse stimulation would also allow implementation of automated methods for DBS programming based on a composite of cortical and peripheral evoked markers [90, 91].

### 4.5. DLEP are better correlated to clinical efficacy than spectral estimates

Following DBS implantation, DBS devices need to be programmed to achieve therapeutic benefit by determining which parameters provide the most benefit[92], through a time-consuming trial-and-error process that depends on clinician’s expertise[92, 93]. A physiological marker that can enable an objective guided surgical targeting and post-operative programming is therefore highly desirable. Prior studies did not compare DLEP to beta power [23, 26], or used only wide-spaced electrodes (1.5mm)[89]. Our results show that DLEP may be more informative than beta power in for selecting optimal contact in the clinic. Interestingly, the aperiodic component of the spectra showed high correlations with clinical efficacy, again highlighting a possible spatial relationship with DLEP.

### 4.6. Limitations

Our study had several limitations: 1) The MER-based localization is constrained to the chosen clinical trajectories, limiting STN sampling (ventral/dorsal STN). 2) Recordings ventral to the STN are sparse, as standard of care does not typically place DBS leads below the STN border. 3) We could not estimate the HFO central frequency across all patients, hence we relied on predefined band based on previous electrophysiological studies[53]. 4) Evaluation of clinical outcomes to derive the clinical efficacy were extracted from program notes of multiple neurologists, possibly introducing bias but increasing generalizability.

## 5. Conclusion

We show how DLEP provide higher spatial specificity for STN-mapping and correlate better with clinical outcomes in Parkinson’s disease patients than beta oscillations, across multiple modalities including intraoperative MER-mapping, post-operative imaging, and post-operative programming. To enable clinical translation of the novel DLEP biomarker, we developed a novel artifact-removal method for monopolar recordings of DLEP, developed and validated a novel method for recovering the amplitude of DLEP at stimulating contacts, and showed that low-frequency stimulation is sufficient for DLEP-based mapping, preserving spatial information without requiring high-frequency bursts. We establish that DLEP amplitude is strongly correlated with aperiodic component of the spectra, which in turn has been shown to correlate with underlying STN excitability, tied to entrainable GPe-STN circuits. This study further highlights that DLEP are a robust and clinically valuable marker for DBS targeting and programming.

## Acknowledgement

The authors thank the subjects for their time and participation. We thank the support from APDA (Post Doctoral Fellowship, Research Grant) and NIH/NINDS (K23 NS097576; R01 NS125143, T32 NS115724).

The authors declare the following financial interests/personal relationships which may be considered as potential competing interests: Enrico Opri reports financial support was provided by National Institute of Neurological Disorders and Stroke and American Parkinson Disease Association. Svjetlana Miocinovic reports financial support was provided by National Institute of Neurological Disorders and Stroke. Jon T Willie reports a relationship with Medtronic Inc that includes: consulting or advisory. Jon T Willie reports a relationship with Abbott Laboratories Inc that includes: consulting or advisory and funding grant. Robert E Gross reports a relationship with NeuroOne Medical, Kriya Therapeutics, Bayer Healthcare, Bluerock Therapeutics, Asklepios BioPharmaceuticals, Aspen Neuroscience, Treefrog Therapeutics, Iota Biosciences, Neuralink that includes: consulting or advisory. Robert E Gross reports a relationship with Medtronic Inc, Boston Scientific Corporation, NeuroPace Inc, Abbott Laboratories Inc that includes: consulting or advisory and funding grant. Robert E Gross reports a relationship with Nia Therapeutics that includes: equity and stock ownership. Nicholas Au Yong’s spouse reports a relationship with Medtronic Inc that includes: equity and stock ownership. Enrico Opri, Svjetlana Miocinovic, Robert E Gross have patent ##18/819,710 pending to Emory University, Georgia Institute of Technology. Enrico Opri, Svjetlana Miocinovic, Jon T Willie have patent #US20240207617A1 pending to Washington University in St. Luis, The Regents of the University of Michigan, Emory University. Svjetlana Miocinovic has patent #US 11,712,564 issued to Emory University. If there are other authors, they declare that they have no known competing financial interests or personal relationships that could have appeared to influence the work reported in this paper.

## Data Availability Statement

The data that support the findings of this study are available upon reasonable request from the corresponding author. The data are not publicly available due to privacy or ethical restrictions.

