## Supplementary Material for "Deep brain stimulation-induced local evoked potentials outperform spectral features in spatial and clinical STN mapping"

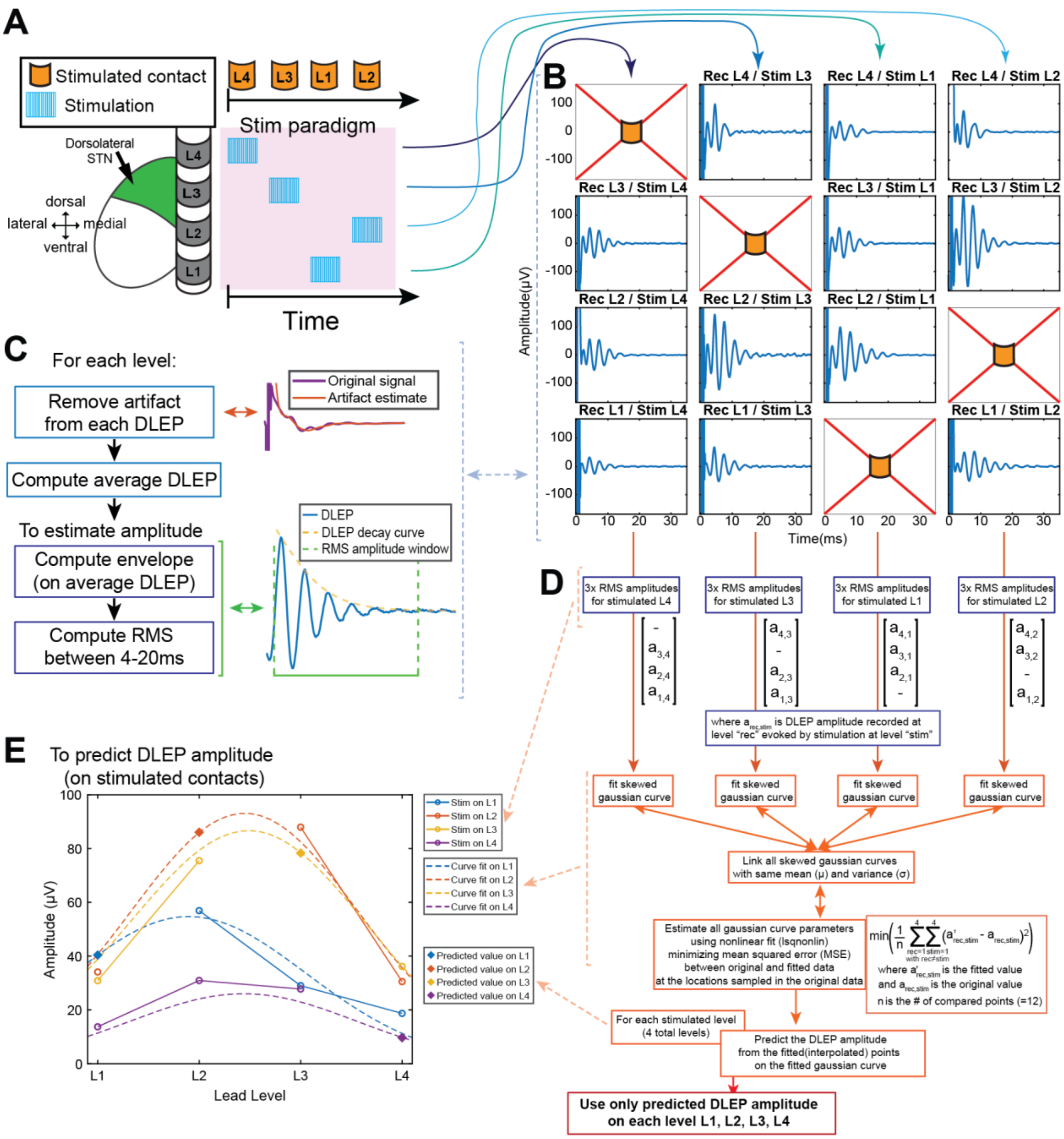


**Supplementary Figure 1. Workflow for estimating DLEP amplitudes on stimulated contacts using a family of skewed normal (Gaussian) distributions.** (A) Stimulation paradigm, illustrating the sequence of stimulated contacts used for acquiring the necessary evoked responses. Only one contact is stimulated at a time along the DBS lead levels (L1–L4). (B) Following each stimulation block, for each contact being stimulated we obtain evoked activity from each non-stimulated contact. In the example here we show the DBS local evoked potential (DLEP) activity recovered after removing the stimulation artifact. Each column represents the data that was recoverable during stimulation of one contact/level (e.g. L4). As each DBS lead has 4 contacts, for every contact stimulated we obtain evoked activity only from the other 3 non-stimulated contacts. (C) As indicated by the blue dashed arrow, the data is used to obtain the necessary DLEP root mean square (RMS) amplitude. Here is the summary of the paradigm used to obtain the RMS amplitude, after removing the stimulation artifact from each evoked activity after stimulation. DLEP RMS is computed between 4-20ms based on the envelope of the obtained DLEP waveform. The envelope is computed with a Hilbert FIR filter. (D) After this step, we obtain 3 average DLEP RMS amplitudes for each stimulated contact ($a_{rec,stim}$), for a total of 12 amplitudes. $a_{rec,stim}$ is the average DLEP RMS amplitude recorded at each level “rec” evoked by stimulation at level “stim”. We then use 4 sets of skewed gaussian curves, one for each amplitude set evoked by the corresponding stimulated contact, linking them via shared mean (μ) and variance (σ) while allowing independent skewness (α). The main text has further details on the mathematical connection between each curve parameters. We leverage a nonlinear fit to fit the appropriate curve parameters that minimize the mean squared error (MSE) between the interpolated DLEP amplitude values from the skewed gaussian curves (indicated as $a'_{rec,stim}$) and the original DLEP amplitude values $a_{rec,stim}$. We perform this minimization including only the “rec” and “stim” combinations that were collected (e.g. “rec”≠“stim”). The fitted curves are then used to interpolate the missing DLEP amplitudes at the stimulation sites (i.e. when “rec”=“stim”). The now predicted DLEP amplitudes on the stimulated contacts are subsequently used in further analysis. (E) Example of skewed gaussian fit across DLEP amplitudes, together with arrows indicating data flow from steps described in (D). Original data is shown with circles, while the predicted (fitted) DLEP amplitudes are shown with diamonds. Dashed lines show the fitted skewed gaussian after the nonlinear fit of the curves’ parameters.

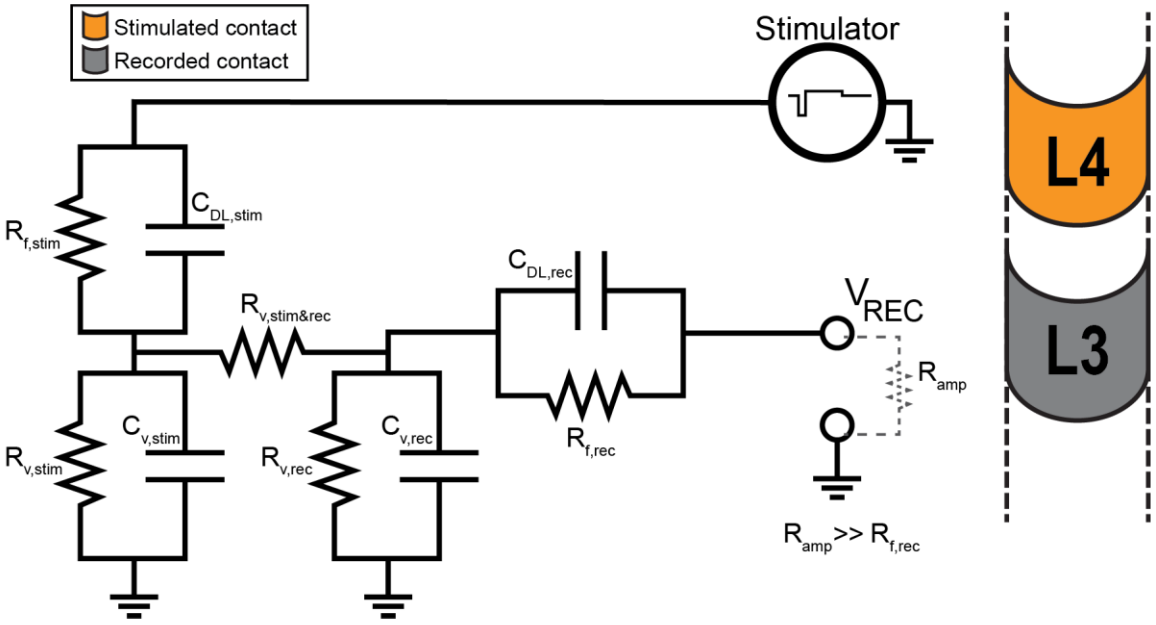


**Supplementary Figure 2. Equivalent circuit-model of the tissue electrode interface between stimulating and recording contact.** This equivalent circuit model was used to design an appropriate artifact estimate to recover the physiology of interest (evoked potential). The current configuration models one contact used for monopolar stimulation (e.g. L4 on a DBS lead), one contact used for monopolar recording (e.g. L3 on a DBS lead), and a return electrode set as ground (the shoulder patch). This model is the same regardless of each pairwise selection of recording and stimulation contacts. Similarly to previous work by Grill group(Kent and Grill, 2012), the electrode-tissue interface included a charge transfer Faradaic resistance $R_{f,stim}$ and $R_{f,rec}$ for the stimulation and recording path, respectively, which were parallel to a double-layer capacitance $C_{DL,stim}$ and $C_{DL,rec}$ for the stimulation and recording path, respectively. The volume conductor included contributions from both the tissue electrode interface of the shoulder patch (parallel $R_{v,stim}$, $C_{v,stim}$ and $R_{v,rec}$, $C_{v,rec}$, for stimulating and recording contact respectively), and the interactions between the stimulating and recording contact ($R_{v,stim\&rec}$). The recorded voltage at the recording contact (e.g. L3) is represented by ($V_{REC}$). The impedance of the amplifier is here shown as $R_{amp}$, however it was not used to model the artifact induced by stimulation. As it is typical in recording setups, $R_{amp}$ is assumed to be much larger than $R_{f,rec.}$ This leads to having a negligible contribution to the recorded $V_{REC}$ by the parallel $R_{f,rec}$ and $C_{DL,rec}$ due to voltage divider effect with $R_{amp}$.

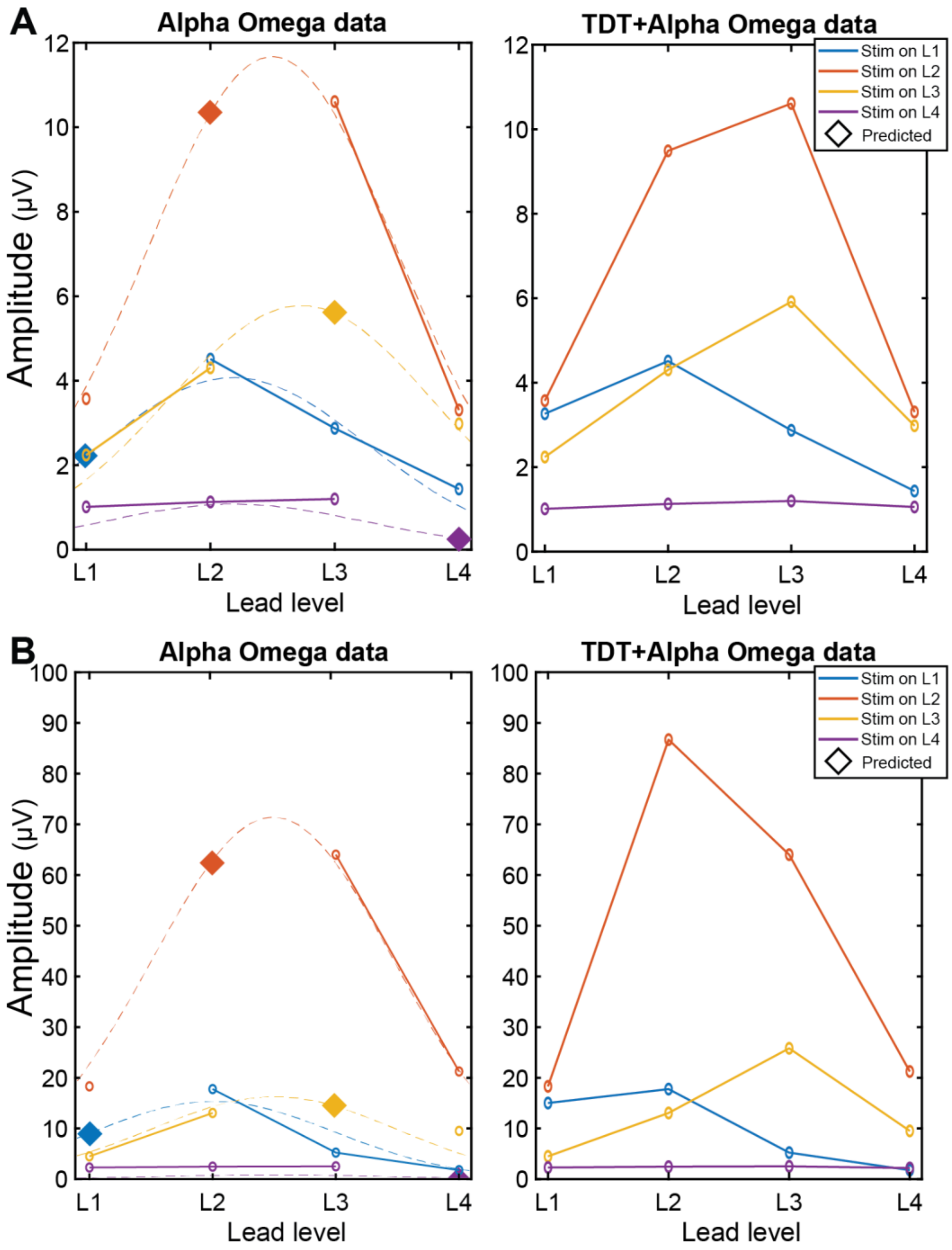


**Supplementary Figure 3. Example of model-predicted DLEP amplitudes at stimulated contact with recorded DLEP in a single patient.** A mathematical model is used to fit gaussian curves to the distribution of DLEP responses and estimate the DLEP amplitude at the stimulated contact (light blue line). We validated the model-based fitting through additional hardware (TDT) capable of recording from the stimulated contacts in a subset of patients. With the clinical system (Neuro Omega), the channel stimulated is not recoverable due to saturation. We tested the recovery of the DLEP at the stimulated contact for both (A) low frequency stimulation and (B) burst of high frequency stimulation (130Hz). Performance across patients is available in Figure 2.

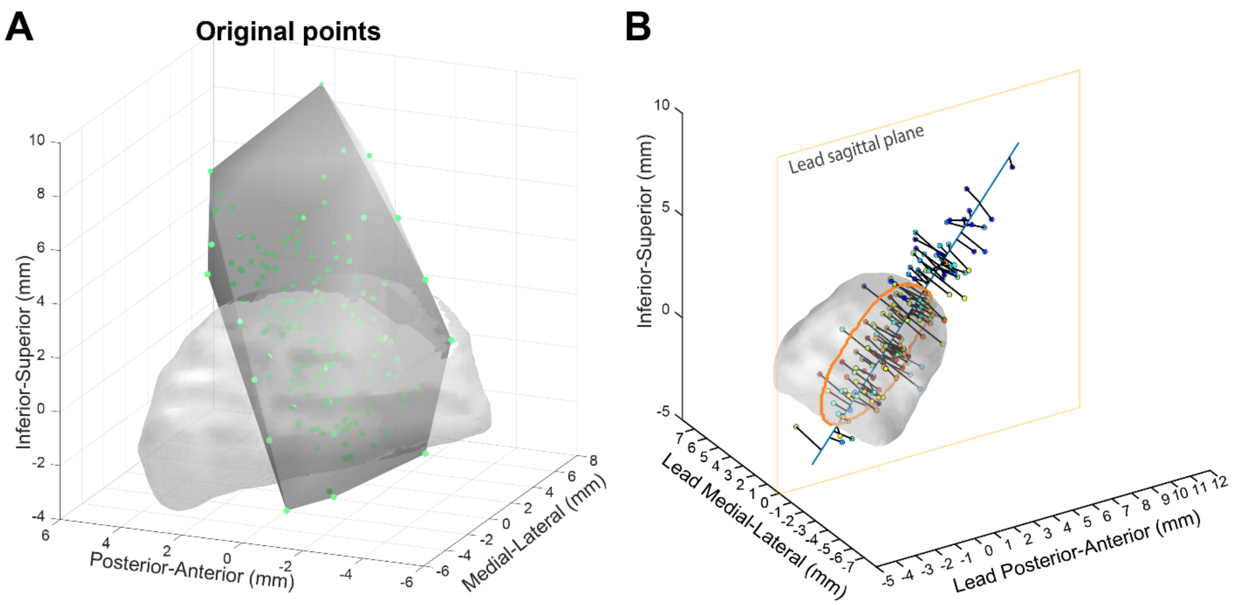


**Supplementary Figure 4. Distribution of DBS contacts localized in MNI space.** (A) Green points represent all DBS contact locations sampled in this study. The light gray volume is STN from MNI atlas. The dark gray volume represents the boundary box for the scattered interpolant (scatteredInterpolant function in MATLAB). (B) To embed the distribution of electrophysiological measures within STN borders for 1D mapping (dorsal to ventral), we obtained the average trajectory (light blue line across STN) across all lead contacts, and projected each electrophysiological measure on the average trajectory itself (to a location orthogonal to the average trajectory from each identified contact; black line towards the average trajectory in light blue). The average trajectory was also used to find the plane orthogonal to the axial plane that contained the average trajectory (lead coronal plane; not shown). This allows us to better visualize a distribution of electrophysiological measures (e.g. DLEP amplitude) across contacts. The objects in the image (contact locations and STN) are rotated to have the new axes aligned with the lead sagittal plane (orange contour represents STN sliced by the lead sagittal plane).

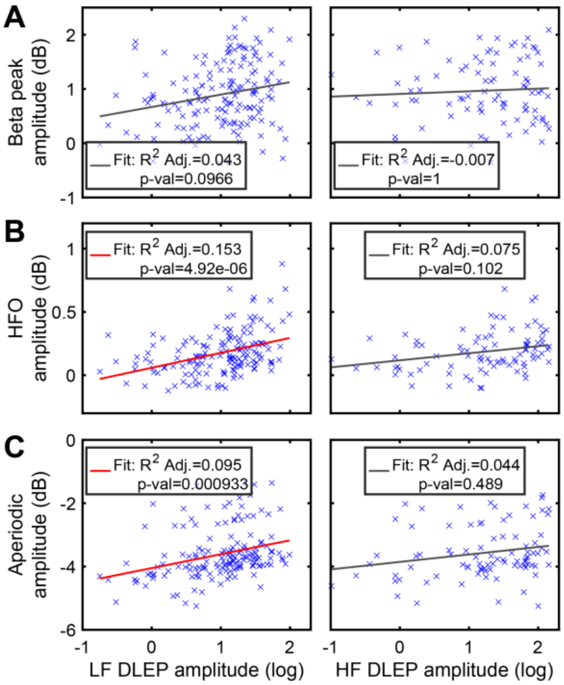


**Supplementary Figure 5. Relationship between absolute DLEP amplitude and spectral features of resting-state LFP signal.** We compared absolute DLEP amplitude (not-z-normalized) across patients with spectral components obtained from resting-state (no stimulation) LFP signals: (A) beta, (B) high frequency oscillations (HFO), (C) high frequency aperiodic component. Left column is LF evoked DLEP, right column is HF evoked DLEP, for each subpanel. There was a weak but significant (B left, LF DLEP R-Adjusted$^2$=0.153, p-val<0.001) relationship between LF DLEP amplitude and HFO amplitude. While beta (A left) and aperiodic component (C left) show a significant trend, the effect explained by their interaction is negligible (R-Adjusted$^2$≤0.095). HF DLEP amplitude shows a significant relationship with HFO amplitude (B right, HF DLEP R-Adjusted$^2$=0.075, p-val<0.01), but the effect explained by their interaction is negligible. HF DLEP does not show any significant trend with other spectral component. This suggests that HFO presence may influence the detectability and manifestation of DLEPs with a larger effect when there is no compounding effect induced by stimulation busts (during LF stimulation).

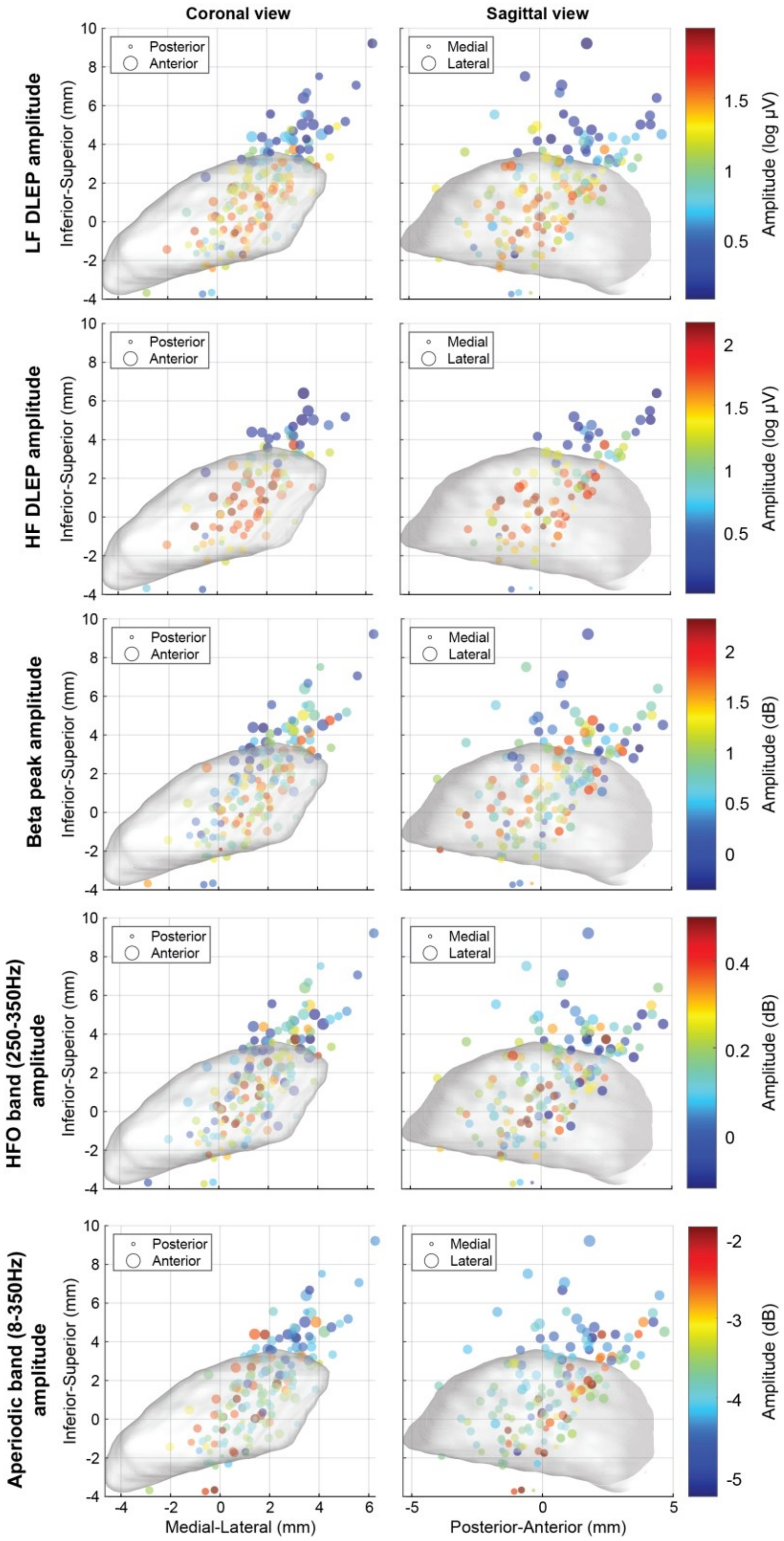


**Supplemental Figure 6. Spatial distribution of DLEP and spectral features in 3D,** in reference to the STN center in MNI space. Showing points from the coronal and sagittal view. Each row shows a different amplitude estimate marker, in order: LF DLEP, HF DLEP, Beta power, HFO power, power of the aperiodic component of the spectra. All points represent the amplitude of the estimated signal at the location of the lead contact where it was sampled, across all the patients.
